# Estimating the parameters of SIR model of COVID-19 cases in India during lock down periods

**DOI:** 10.1101/2020.06.03.20120899

**Authors:** Dilip Kumar Bagal, Arati Rath, Abhishek Barua, Dulu Patnaik

## Abstract

From the pandemic scenario of COVID-19 disease cases in all over the world, the outbreak prediction becomes very complex for the emerging scientifically research. Several epidemiological mathematical models of spread are increasing day by day to forecast correctly. Here, the classical SIR modelling approach is carried out to study the different parameters of this model in case of India county. This type of approach analyzed by considering different governmental lock down measures in India. There are some assumptions were taken into account for fitting the model in Python simulation in each lock down scenario. The predicted parameters of SIR model showed some improvement in each case of lock down in India. The outcome results showed the extreme interventions should be taken to tackle this type of pandemic situation in near future.

**Author approval:** This article does not contain any studies with human participants or animals performed by any of the authors.

**Competing interests:** The authors declare no competing interests.

**Declarations:** The authors declare that there is no conflict of interest regarding the publication of this article.

**Data availability statement:** The working data set used for this study has been submitted to the journal as additional supporting files.

**Data availability links:**

1. https://github.com/CSSEGISandData/COVID-19/tree/master/csse_covid_19_data/csse_covid_19_time_series
2. https://www.dropbox.com/sh/akc525jjq3dp485/AADgo6WsT1RBpZqahmj_k-v_a/SIR/italy_fit.py?dl=0.

## Introduction

On 3rd December 2019, a new virus came into the picture with 41 patients connected mysterious pneumonia cases in the big seafood market area of Wuhan city, China country [1]. At that time, this virus was called SARS-CoV version 2 and after WHO named it as COVID-19 with global pandemic on 11th March 2020 [2]. COVID-19 is the infectious disease caused by the severe acute respiratory syndrome novel coronavirus (SARS-CoV-2) [3]. Now, almost all the parts of the world are exposed to this virus and since the infectibility of this virus is large, a huge number of people have already become victim of it [4]. SARS-CoV-2 infection has represented the one of the largest challenges for humanity. India first reported a COVID-19 case in a student who returned from Wuhan, the capital Hubei province, China on January 30, 2020 [5]. Figure 1 showed the COVID-19 scenario of World till 31st May 2020 by considering conformed, deaths and recovered cases.

**Figure 1:**
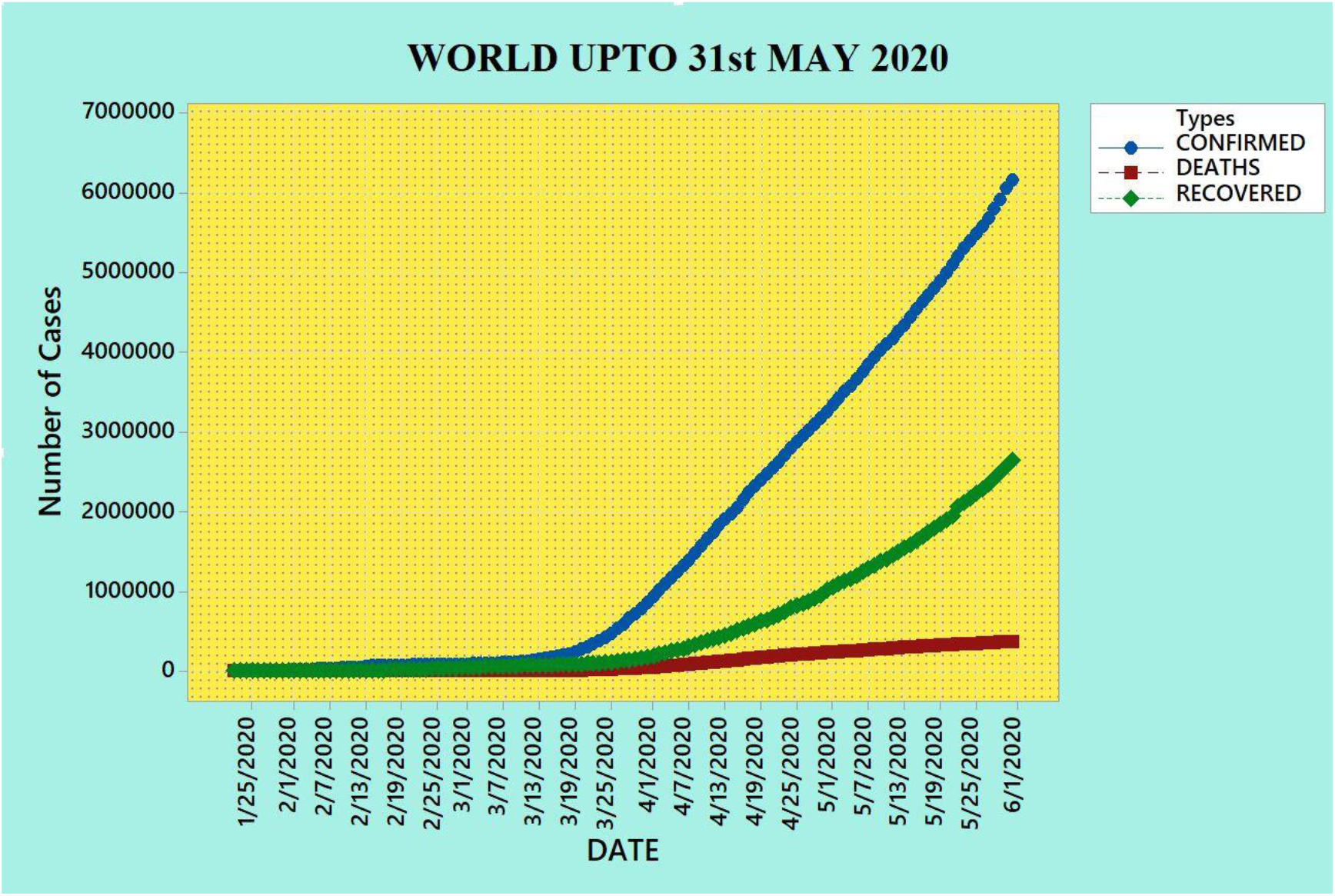
Scenario of our World till 31^st^ May 2020

Dhanwant and Ramanathan [6] implemented the Susceptible-Infected-Recovered (SIR) approach for forecasting the outbreaks of COVID-19 cases in India using Scipy platform. Xavier et al. [7] proposed an epidemiological model combined SIR to a genetic algorithm with three types of networks. Elhia et al. [8] optimized the SIR epidemic model with time variation and controlled measures of H1N1 data in Morocco. Here five parameters such as recruitment rate of susceptible, effective contact rate, natural mortality rate, recovery rate and disease induced death rate had been taken into account for modelling. Chaves et al. [9] determined the reproduction number (R_0_) and time-varying reproduction number (R_t_) for Costa Rica, Panama and Uruguay from the SIR model with three types of phase. Singh and Adhikari [10] studied the age-structured impact of India, China and Italy using hybrid SIR model. Das [11] predict the COVID-19 disease progression in India and China using SIR and Statistical Machine Learning approach by taking data set till 6^th^ April 2020. Arif et al. [12] analyzed the COVID-19 data set up to 10^th^ May 2020 using SIR model for Pakistan country. Boudrioua et al. [13] predicted the daily infected cases with COVID-19 in Algeria using SIR approach with the data set ranging from 25^th^ February 2020 to 24^th^ April 2020. Deo et al. [14] predicted the dynamics of COVID-19 epidemic data set from 2^nd^ March 2020 to 30^th^ April 2020.in India based on time-series SIR model. Hazem et al. [15] analyzed the COVID-19 cases in United States, Germany, the United Kingdom, Italy, Spain, and Canada through SIR approach from 20^th^ January 2020 to 15^th^ May 2020 considering lockdown measures. Jakhar et al. [16] predicted COVID-19 epidemic cases in 24 different States of India using SIR Model by taking data set up to 12^th^ May 2020. Mujallad and Khoj [17] forecasted COVID-19 cases using the SIR Model in the city of Makkah, Saudi Arabia taking into account the dataset from 16^th^ March 2020 to 9^th^ May 2020. Oliveira et al. [18] forecasted by introducing a Bayesian methodology to the SIR model COVID-19 cases in Brazil using dataset 26^th^ February 2020 to 20^th^ May 2020. Postnikov [19] analyzed SIR model with the Verhulst (logistic) equation approach for the COVID-19 outbreak in different countries using MATLAB software. Gallardo et al. [20] analyzed the Phylodynamic SIR trajectories of COVID-19 cases in Latin America using data set up to 13^th^ May 2020 with two epidemiological surveillances.

Lopez and Rodo [21] predicted the outbreaks in Spain and Italy through the Susceptible-Exposed-Infectious-Removed (SEIR) model till 31^st^ march 2020 with multiple scenarios. Yang et al. [22] studied the modified SEIR and Artificial Intelligence (AI) approach for the prediction of COVID-19 outbreaks under certain public interventions in China using time slice data. Engbert et al. [23] predicted the dynamics of COVID-19 cases in Germany through stochastic SEIR model with different scenarios. Berger et al. [24] studied the role of testing the samples and case-dependent quarantine measures in United States through SEIR epidemic model. Hou et al. [25] studied the well mixed approach of SEIR model of COVID-19 cases in Wuhan city of China with suggestion of interventions such as quarantine and isolation. Saito et al. [26] applied the SEIR model on the cases of H1N1 influenza pandemic in Japan with detection of the immigration effect. Godio et al. [27] implemented a generalized SEIR epidemiological model to Italian population of SARS-CoV-2 cases by comparing with Spain and South Korea through Matlab software. Feng et al. [28] studied the COVID-19 outbreak using SEIR model by enacting strict social distancing policies to slow down the spread of virus. Gupta et al. [29] studied the SEIR and regression approach on COVID-19 cases in India by training data up to 30^th^ March 2020. Pandey et al. [30] predicted the COVID-19 cases in India using SEIR and Regression model from 30^th^ January 2020 to 30^th^ March 2020 with the determination of R_0_ value. Goswami et al. [31] emphasized on the effective contact rate of COVID-19 cases in India by applying SEIR model and compared with other six countries. Bonnasse-Gahot et al. [32] used the SEIR type model with incubation compartment in France and also emphasized on the monitoring od bed availability during pandemic. Dixit et al. [33] forecasting using Regression, exponential smoothing and compartmental age structured SEIR modelling of COVID-19 cases up to 25^th^ May 2020 of India. Kohanovski et al. [34] fitted an SEIR model to COVID-19 cases using data from 12 countries along with extreme non-pharmaceutical interventions (NPIs). Teles [35] studied the SARS-COVID-2 epidemic outbreak cases through time-dependent SEIR model in Portugal. Wagh et al. [36] studied the epidemic peak for COVID-19 cases in India utilizing Auto-Regressive Integrated Moving Average (AIRMA) model coupled with SEIR Compartmental epidemiological model till 9^th^ May 2020 dataset.

Ray et al. [37] studied the short-and long-term impact of an initial 21-day lockdown on March 25 through eSIR model approach considering the dataset from 1^st^ March 2020 to 7^th^ April 2020 by RShiny package. da Cruz et al. [38] analyzed the COVID-19 outbreak in the State of São Paulo, Brazil using SEIR-A model. Kobayashi et al. [39] studied the intervention effect in Japan for COVID-19 cases taking data up 18^th^ May 2020 using SS-SIR model approach. Leon et al. [2] forecasted the outbreak of Maxico through SEIARD model with the determination of R_0_ value using Matlab 2016b software which based on Runge-Kutta formula. Rajesh et al. [40] analyzed the COVID-19 data set of India from 22^nd^ March 2020 to 17^th^ April 2020 using SIR(D) dynamical model in India. Khatua et al. [3] gave a dynamic optimal SEIAR model for COVID-19 cases in India along with sensitivity analysis and recommended that consecutive 40 days might be able to control the pandemic influence. Jiwei et al. [41] studied the SEIRQAD model with control stages of different regions of China with some meteorological influences. Chatterjee et al. [42] applied the stochastic SEIQRD model of COVID-19 cases in India by taking into account data of 30^th^ March 2020. Singh et al. [43] predict the COVID-19 data set of India and its states till 12th May 2020 using a mathematical approach. Hao [44] used a MSIR and MSEIR model to predict the outbreak of COVID cases from 21st January 2020 to 17^th^ February 2020 in China. Mandal et al. [45] analyzed the cases of COVID-19 in three states of India by five time-dependent classes i.e. Susceptible S(t), Exposed E(t), Hospitalized infected I(t), Quarantine Q(t) and Recovered or Removed R(t). Khan and Hossain [46] made an empirical analysis on COVID-19 outbreaks in Bangladesh by projection technique called Infection Trajectory-Pathway Strategy (ITPS).

Pinter et al. [47] implemented the hybrid machine learning approach to predict the outbreaks of COVID-19 cases in Hungary using data set from 4^th^ March 2020 to 19^th^ April 2020. Villaverde and Jones [48] estimated the parameters using a standard epidemiological SIRDC model of COVID-19 in many Countries, States, and Cities. Beck [49] studied a modified confinement compartment SEICRS model for COVID-19 dataset from 3^rd^ February 2020 to 14^th^ May 2020 in Costa Rica Mathematica 12 software with governmental non-pharmaceutical interventions (NPIs). Gupta [50] forecasted the COVID-19 cases for India, Italy, USA and UK countries till 18^th^ May 2020 data set. Lyra et al. [51] modelled the COVID-19 cases with SEIR(+CAQH) approach in Brazil and found fatality rate of disease appears to be higher among the elderly population (particularly 60 and beyond). Rustam et al. [52] forecasted COVID-19 cases by four machine learning regression models i.e. (i) Linear Regression, (ii) LASSO Regression, (iii) Support Vector Machine and (iv) Exponential Smoothing for different countries. Gupta et al. [53] analyzed the COVID-19 outbreaks of India and several states with exponential modeling with consideration of several lock down measures. Now, the Figure 2 showed the COVID-19 scenario of India till 31^st^ May 2020 by considering conformed, deaths and recovered cases.

**Figure 2:**
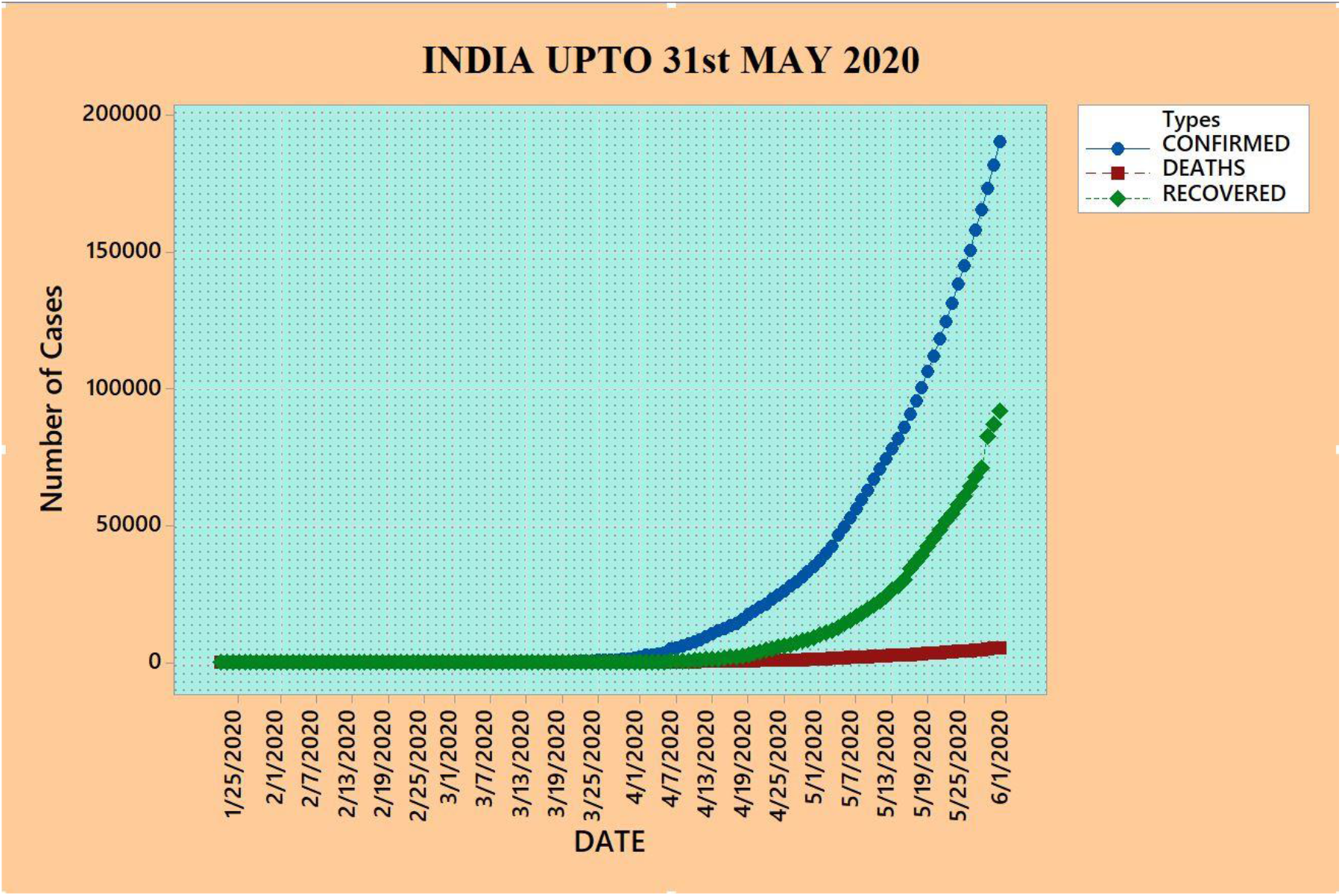
Scenario of India till 31^st^ May 2020

### Estimation of parameters of SIR model for INDIA from actual data set

For the epidemical mathematical model, there were used of basic model based on compartments such as:

a. (Susceptible->Infectible) SI model
b. (Susceptible->Infectible-> Susceptible) SIS model and
c. (Susceptible->Infectible-> Recovery/Removed) SIR model

In 1927, firstly Kermack and McKendrick proposed a class of compartmental models that simplified the mathematical modeling of infectious disease transmission. Entitled as SIR model, it is a set of general equations which explains the dynamics of an infectious disease spreading through a susceptible population.

**Figure 3:**
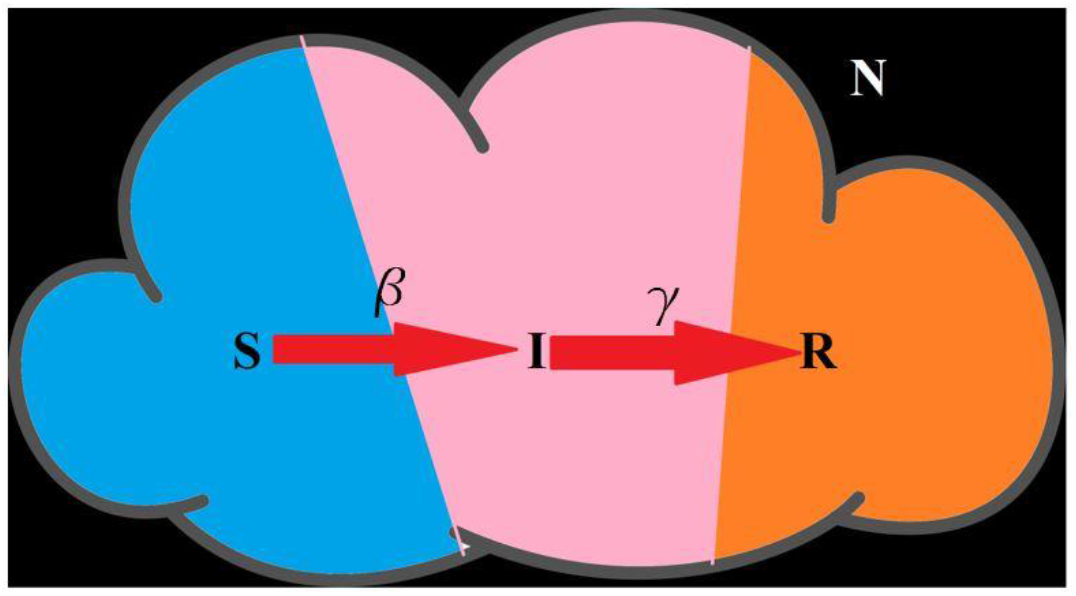
Division of population based on SIR model

Essentially, the standard SIR model is a set of differential equations that can suit the Susceptible (if previously unexposed to the pandemic disease), Infected (if currently colonized by the pandemic disease), and Removed (either by death or recovery) as follows:

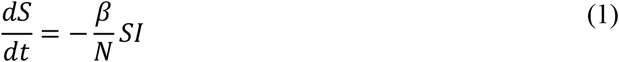

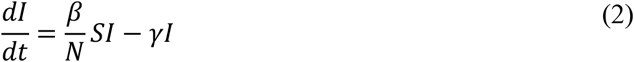

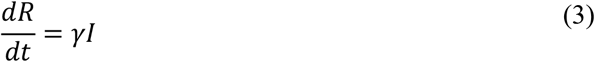

Here, N=S+I+R, is independent of time t, denotes the total population size [8, 22, 54-56]. India 2020 population is estimated at **1,380,004,385** people at mid-year according to UN data. Therefore, the value of population size (N) is considered as 1,380,004,385 for the SIR modelling of India [57].

**Figure 4:**
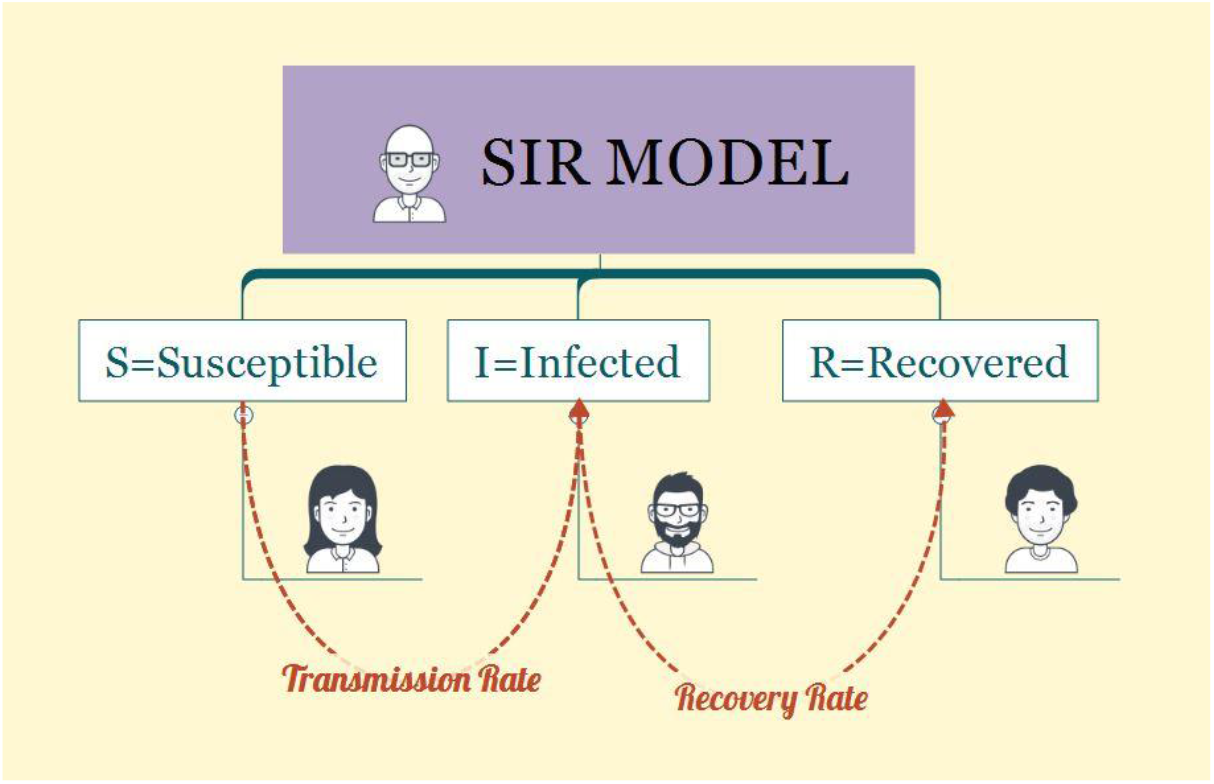
Basic structure of SIR model

Figure 4: Basic structure of SIR model

When there is no infection means I+R=0 then putting S≈N in Equation (2), we get

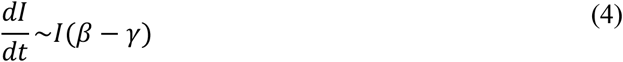

Then,

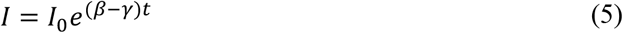

#### Determination of β and m value

At the onset of infection, almost all population is susceptible i.e. S≈N. So, I(t) first grows exponentially.

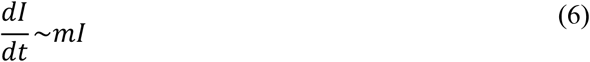

Where, **m**= **β**- **γ**, is a constant term which is the difference between transmission rate and recovery rate.

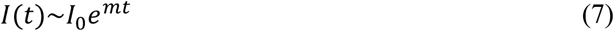

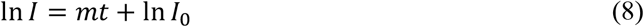

It can be estimate m value by looking at the data on a log-plot and using for example least squares to fit best-line fit.

In this work, the data set was taken from the 2019 Novel Coronavirus Visual Dashboard operated by the Johns Hopkins University Center for Systems Science and Engineering (JHU CSSE) database [58]. And, the Python code [59] of SIR model based on dataset of India was simulated on Google Colab Platform and the estimated futuristic dates are determined through the online date calculator [60]. Also, the scenario of our World and India based on actual JHU-CSSE data set was computed through Minitab software [57].

In this study, we assume the governmental protocols of each lockdown measure is homogeneously implemented across the country. However, because of significant differences in various socio-economic, demographic, cultural, and administrative level factors, actual transmission rates are bound to differ from region to region. Hence, the estimated parameters in our study are only valid for overall predictions of cases in India, on an average, and may fail to trace the dynamics of the epidemic in sub-regions, say districts or states [14]. The uncertainty in our predictions is large due to many unknowns arising from model assumptions, population demographics, the number of COVID-19 diagnostic tests administered per day, testing criteria, accuracy of the test results, and heterogeneity in implementation of different government-initiated interventions and community-level protective measures across the country. We have neither accounted for age-structure, contact patterns or spatial information to finesse our predictions nor considered the possibility of a latent number of true cases, only a fraction of which are ascertained and observed. COVID-19 hotspots in India are not uniformly spread across the country, and state-level forecasts may be more meaningful for state-level policymaking. Regardless of the caveats in our study, our analyses show the impact and necessity of lockdown and of suppressed activity post-lockdown in India. One ideological limitation of considering only the epidemiological perspective of controlling COVID-19 transmission in our model is the inability to count excess deaths due to other causes during this period, or the flexibility to factor in reduction in mortality/morbidity due to some other infectious or flu-like illnesses, traffic accidents or health benefits of reduced air pollution levels. A more expansive framework of a cost-benefit analysis is needed as we gather more data and build an integrated landscape of population attributable risks [51].

**Figure 5:**
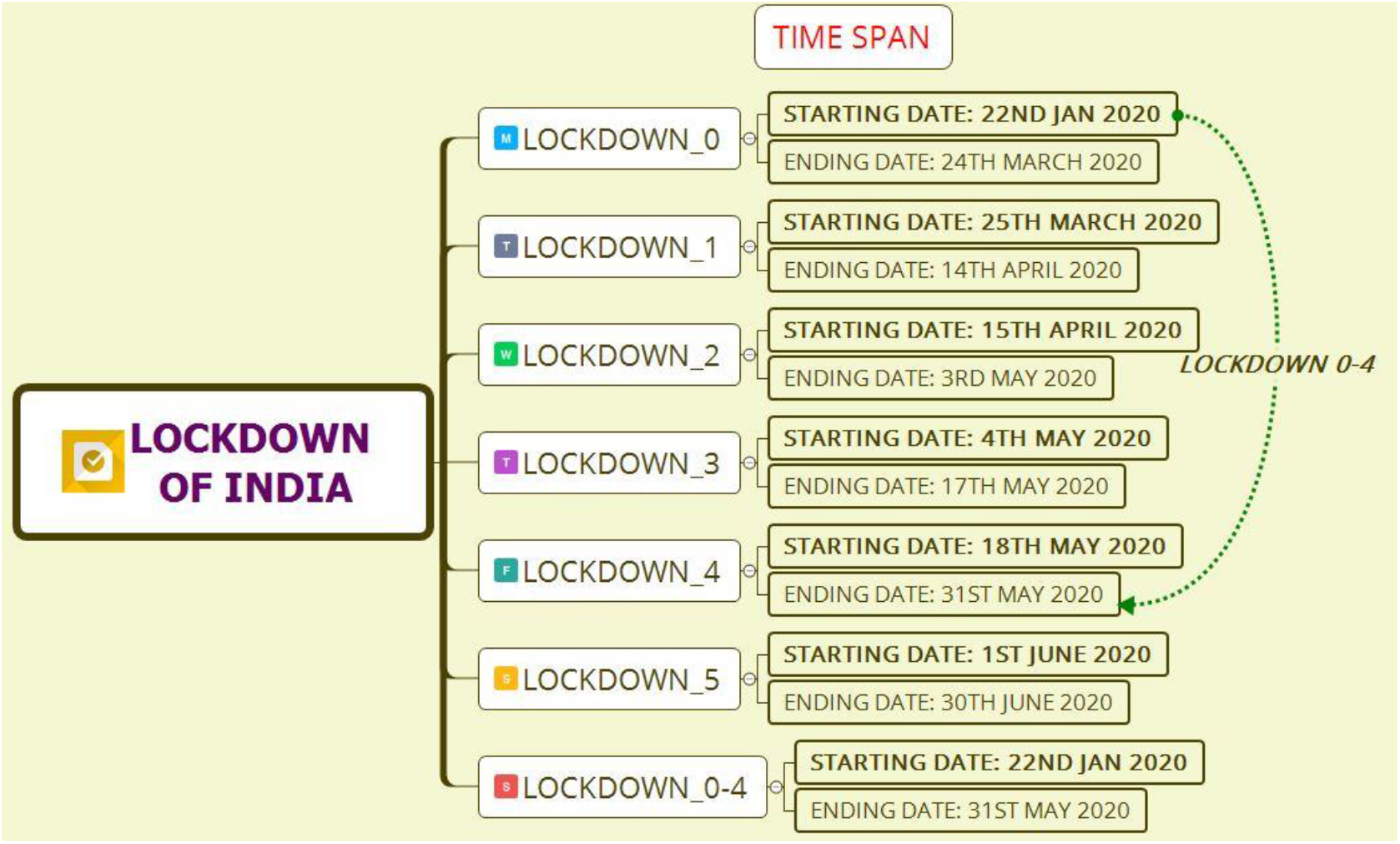
Time span of lock down in India

Using COVID-19 dataset from India, we get:

**Table 1:** Obtained values of average *γ*, *β* and m

| Sl. No. | Lock Down | Starting Date | Ending Date | AVG. ($\gamma$) | AVG. ($\beta$) | $m$ |
| --- | --- | --- | --- | --- | --- | --- |
| 1 | LD_0 | 22nd Jan 2020 | 24th March 2020 | 0.0195558 | 0.240961 | 0.221405 |
| 2 | LD_1 | 25th March 2020 | 14th April 2020 | 0.0263881 | 0.128706 | 0.102318 |
| 3 | LD_2 | 15th April 2020 | 3rd May 2020 | 0.086535 | 0.086535 | 0.051772 |
| 4 | LD_3 | 4th May 2020 | 17th May 2020 | 0.0494598 | 0.0816826 | 0.032222 |
| 5 | LD_4 | 18th May 2020 | 31st May 2020 | 0.0622677 | 0.0850051 | **0.022737** |
| 6 | LD_0-4 | 22nd Jan 2020 | 31st May 2020 | 0.0622677 | 0.0850051 | 0.022737 |

Here, the lock down number four gave the minimum value that means severe control measures were taken to control the spread of virus.

#### Determination of *γ* value

Suppose I(t)=I_0_ (Constant term), then

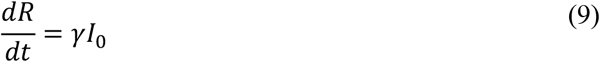

Then,

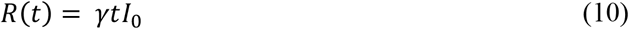

If it takes t=T Days to recover, then R(T)=I_0_, then γT=1.

So,

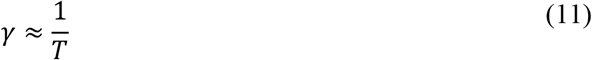

Where, T is the recovery period.

From Equation (3), for change in time of dt=a, then

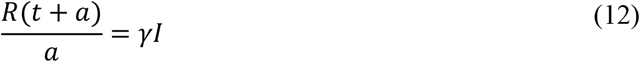

Then,

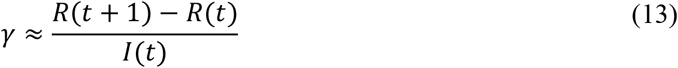

Estimating directly from dataset of India, we obtained:

**Table 2:** Obtained values of average *γ*

| Sl. No. | Lock Down | Starting Date | Ending Date | AVG. ($\gamma$) |
| --- | --- | --- | --- | --- |
| 1 | LD_0 | 22nd Jan 2020 | 24th March 2020 | 0.0195558 |
| 2 | LD_1 | 25th March 2020 | 14th April 2020 | 0.0263881 |
| 3 | LD_2 | 15th April 2020 | 3rd May 2020 | **0.086535** |
| 4 | LD_3 | 4th May 2020 | 17th May 2020 | 0.0494598 |
| 5 | LD_4 | 18th May 2020 | 31st May 2020 | 0.0622677 |
| 6 | LD_0-4 | 22nd Jan 2020 | 31st May 2020 | 0.0622677 |

Here, the lock down number two gave the maximum value that means the recovery rate of population is better than other lock down periods.

#### Determination of I_max_ and S_inf_

Now divide Equation (2) by Equation (1), we obtained:

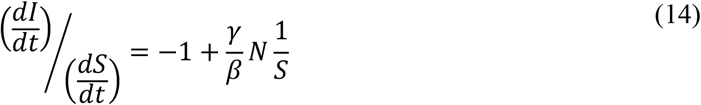

Then,

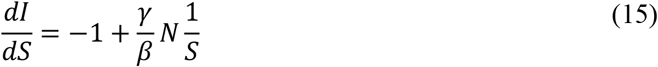

Then integrating both sides, we get:

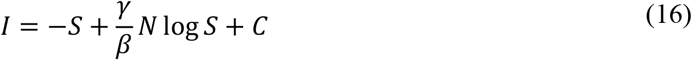

Where C= Constant Term

Now at the onset of infection, I value of Equation (16) is very small whereas S≈N (Total Population size).

So, at t=0, we plug in I~0 and S~N:

Now, from Equation (16),

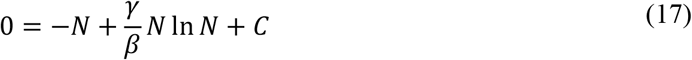

Then,

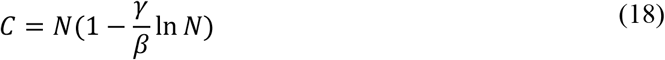

Putting the value of C of Equation (18) in Equation (16), we get

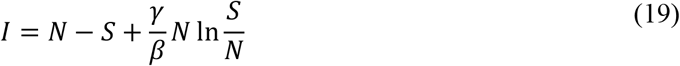

This equation is then valid for all time. In general, I initially grow exponentially, then reaches a peak and then gradually decays back to zero value. Now finding out the percentage of sick people at the peak of infection (I_max_) and susceptible people still remaining after the infection of COVID-19 virus passes. From Equation (2) and (20), which are differential equation and its solution.

Again, to simplify, assume S= N_s_, I= N_i_ and R= N_r_. Where, s, i and r represents the fraction of total susceptible, infected and recovery/removed population. Then,

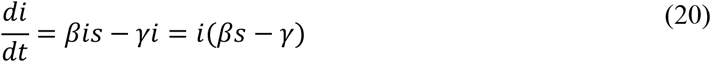

And

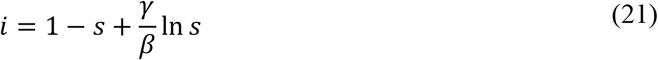

Then, the peak infection happens when di/dt=0 and s value comes to:

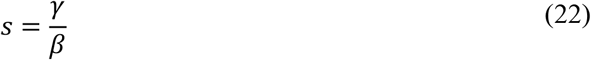

Then plug the Equation (22) into the Equation (21), we obtain:

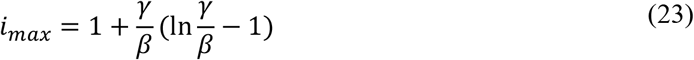

From the dataset of India, we find:

**Table 3:** Obtained values of I max

| Sl. No. | Lock Down | Starting Date | Ending Date | $I_{max}$ (in %) |
| --- | --- | --- | --- | --- |
| 1 | LD_0 | 22nd Jan 2020 | 24th March 2020 | 71.6 |
| 2 | LD_1 | 25th March 2020 | 14th April 2020 | 47.1 |
| 3 | LD_2 | 15th April 2020 | 3rd May 2020 | 23.2 |
| 4 | LD_3 | 4th May 2020 | 17th May 2020 | 9.08 |
| 5 | LD_4 | 18th May 2020 | 31st May 2020 | 3.95 |
| 6 | LD_0-4 | 22nd Jan 2020 | 31st May 2020 | 3.95 |

The percentage of sick people at the peak of infection drastically reduced in each lock down of India. For finding out 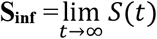 i.e. the percentage of susceptible people still remaining after the infection passes away. We note that i=0, as t tends to infinity (∞) at the end of infection, then the Equation (21) becomes:

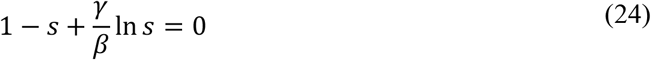

Equation (24) can be solved numerically for s value. Now from the data set of India we got the value of **S_inf_** for every lock down stages.

**Table 4:** Obtained values of S_inf

| Sl. No. | Lock Down | Starting Date | Ending Date | $S_{inf}$ |
| --- | --- | --- | --- | --- |
| 1 | LD_0 | 22nd Jan 2020 | 24th March 2020 | 99.98 |
| 2 | LD_1 | 25th March 2020 | 14th April 2020 | 99.21 |
| 3 | LD_2 | 15th April 2020 | 3rd May 2020 | 89.09 |
| 4 | LD_3 | 4th May 2020 | 17th May 2020 | 66.3 |
| 5 | LD_4 | 18th May 2020 | 31st May 2020 | 45.53 |
| 6 | LD_0-4 | 22nd Jan 2020 | 31st May 2020 | 45.53 |

The percentage of sick people at the end of infection reduced as per the strict procedures of lock down and this can be further decreased for future lock down implementations.

#### Determination of R_0_

The special term of basic reproduction number i.e. R_0_ is the ratio of transmission rate and recovery rate.

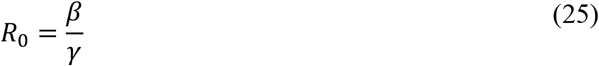

It represents the number of individuals (on average) that a single individual infects [7, 56, 61-68]. In terms of R_0_, we have

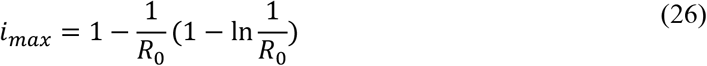

And

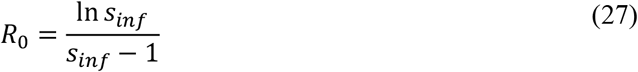

From the dataset of India, we get

**Table 5:**
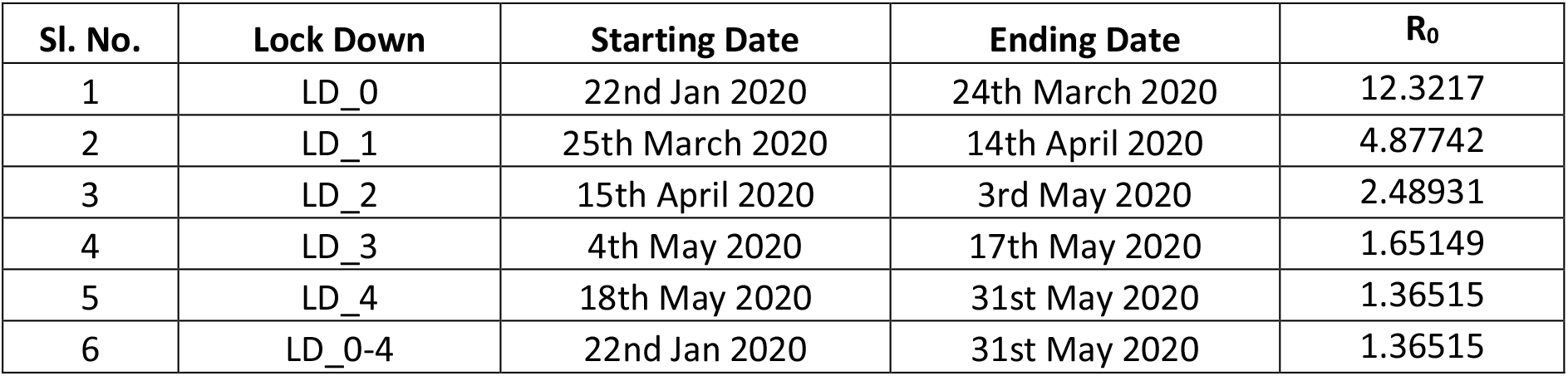
Obtained values of R_0

| Sl. No. | Lock Down | Starting Date | Ending Date | $R_0$ |
| --- | --- | --- | --- | --- |
| 1 | LD_0 | 22nd Jan 2020 | 24th March 2020 | 12.3217 |
| 2 | LD_1 | 25th March 2020 | 14th April 2020 | 4.87742 |
| 3 | LD_2 | 15th April 2020 | 3rd May 2020 | 2.48931 |
| 4 | LD_3 | 4th May 2020 | 17th May 2020 | 1.65149 |
| 5 | LD_4 | 18th May 2020 | 31st May 2020 | 1.36515 |
| 6 | LD_0-4 | 22nd Jan 2020 | 31st May 2020 | 1.36515 |

The basic reproduction number reduced as per the strict procedures of lock down and this can be further decreased. The simulation results of Python code of SIR model based on India COVID-19 dataset was shown in Appendix-I (From Figure 6-11) for each lock down cases.

## Conclusion

From study of SIR model for the case of India, several points come into conclusions and these are described as the betterment of each lock down measures should be taken. The recovery rate is increasing and the transmission rate is decreasing as a result the value of basic reproduction number is decreasing by flatten the curve of epidemic spread of COVID-19. Similarly, the percentage of susceptible people still remaining after the infection passes away decreased in horrible manner and it should be checked near future. Again, the peak percentage value of infectious people after the sickness minimized with better values. In case of India, further strict governmental interventions should be chosen and of course the pandemic scene can be drastically reduced by the heartfully awareness among residing people.

## Data Availability

The working data set used for this study has been submitted to the journal as additional supporting files.

https://github.com/CSSEGISandData/COVID-19/tree/master/csse_covid_19_data/csse_covid_19_time_series

## Funding statement

There is no funding for this study.

## Supplementary material

The Python scripts used to generate this article, which include the code of SIR model used to derive parameters, will be made available with this manuscript.

## Appendix-I

**Figure 6:**
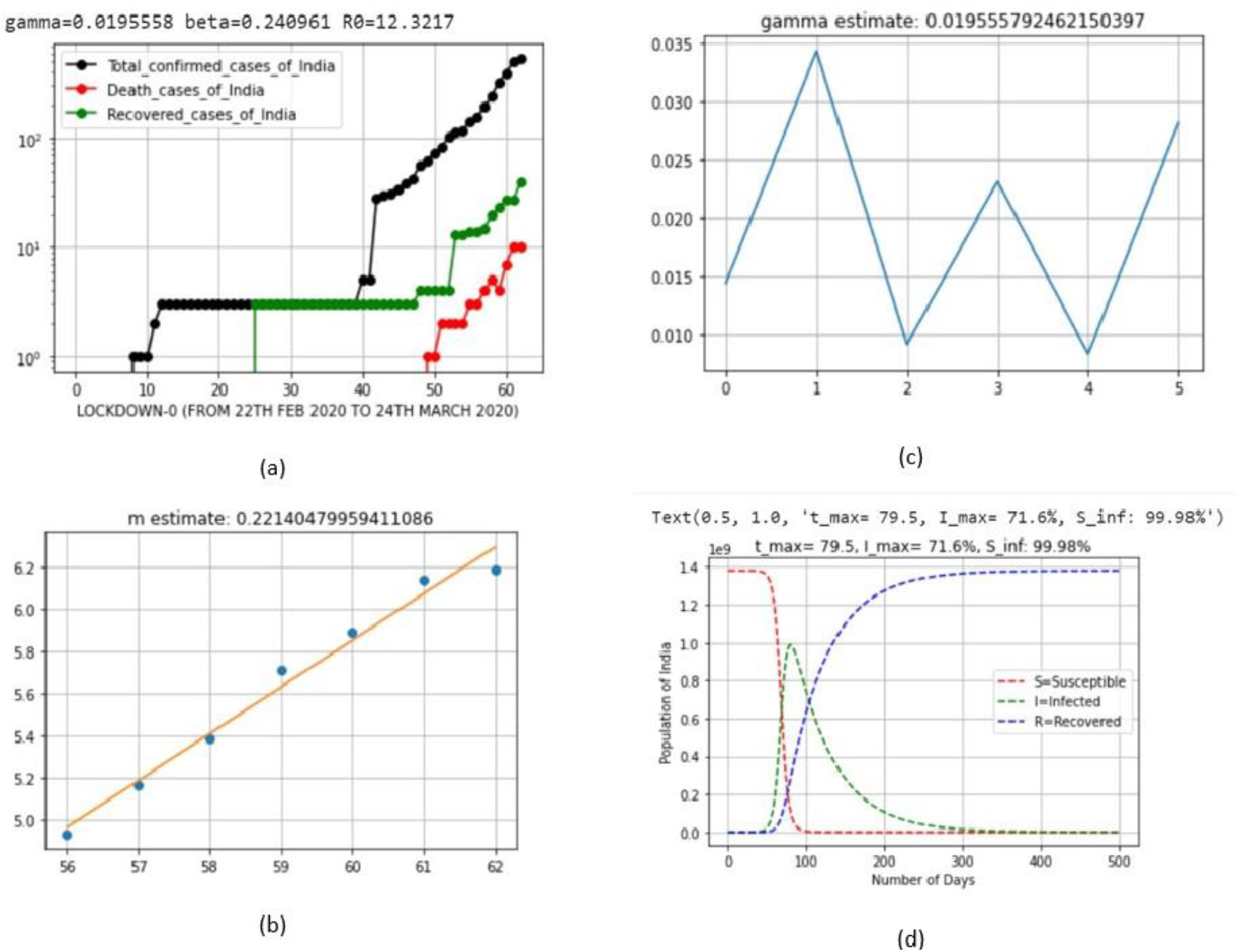
Output parameters of SIR model for lock down 0

**Figure 7:**
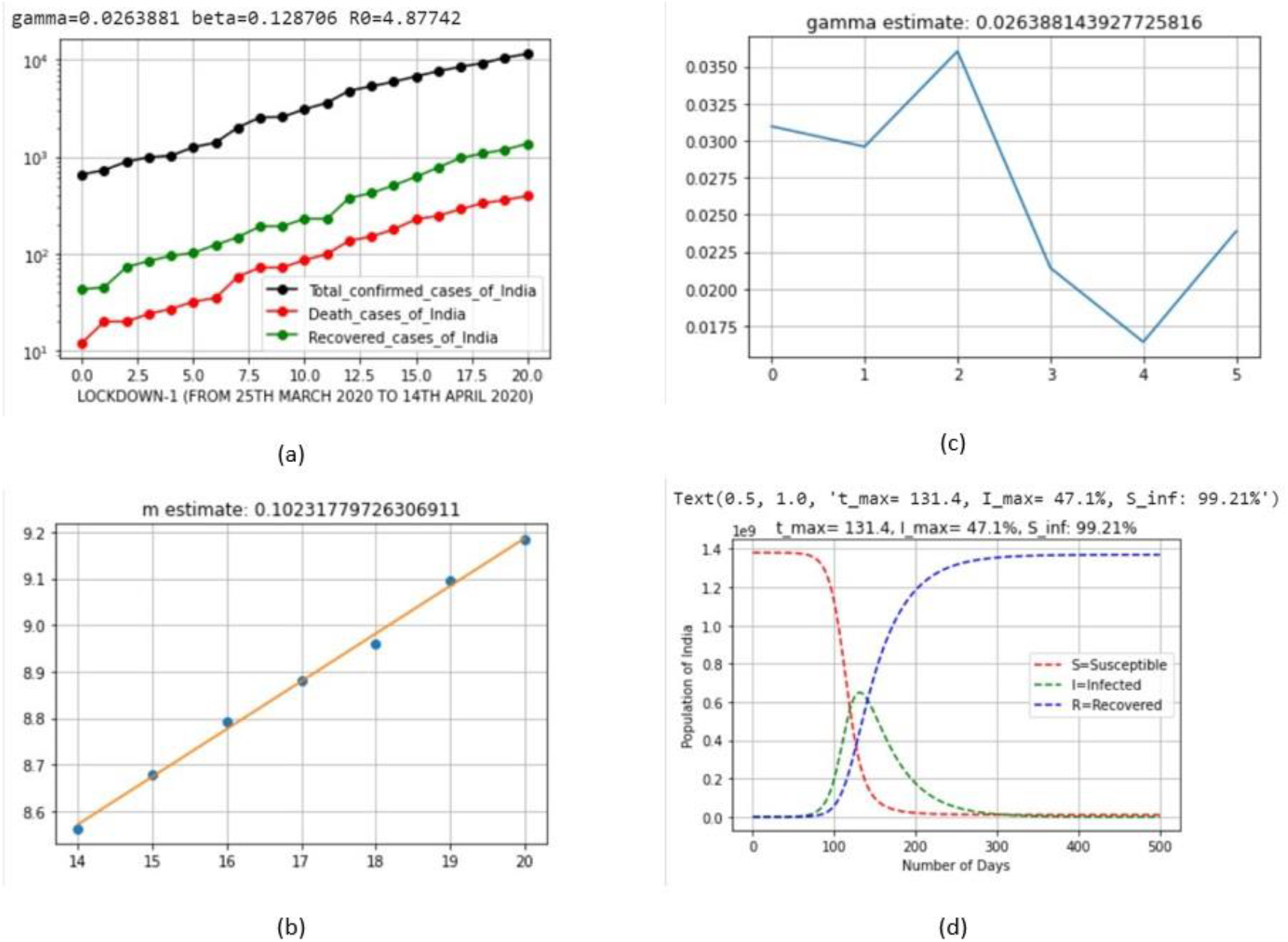
Output parameters of SIR model for lock down 1

**Figure 8:**
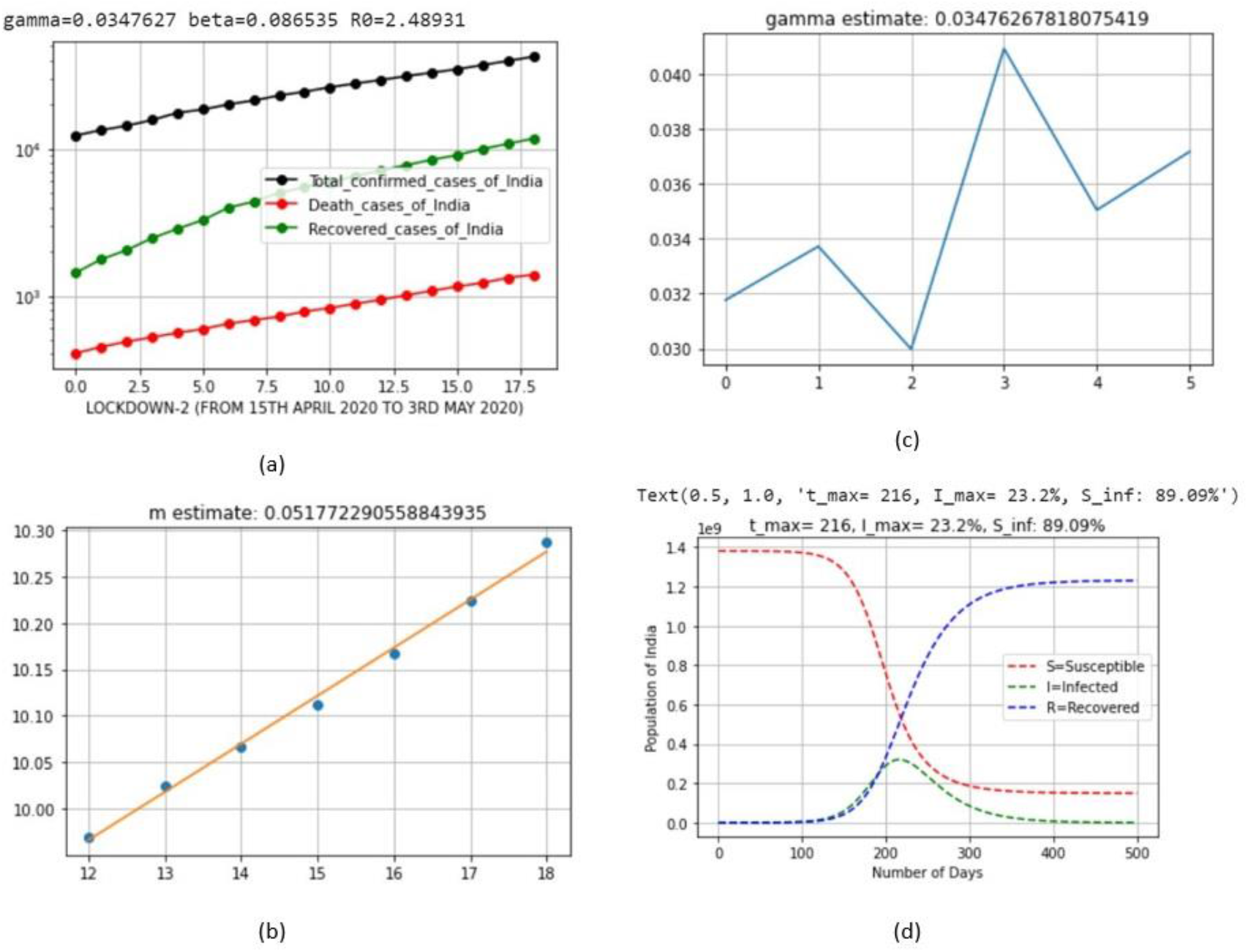
Output parameters of SIR model for lock down 2

**Figure 9:**
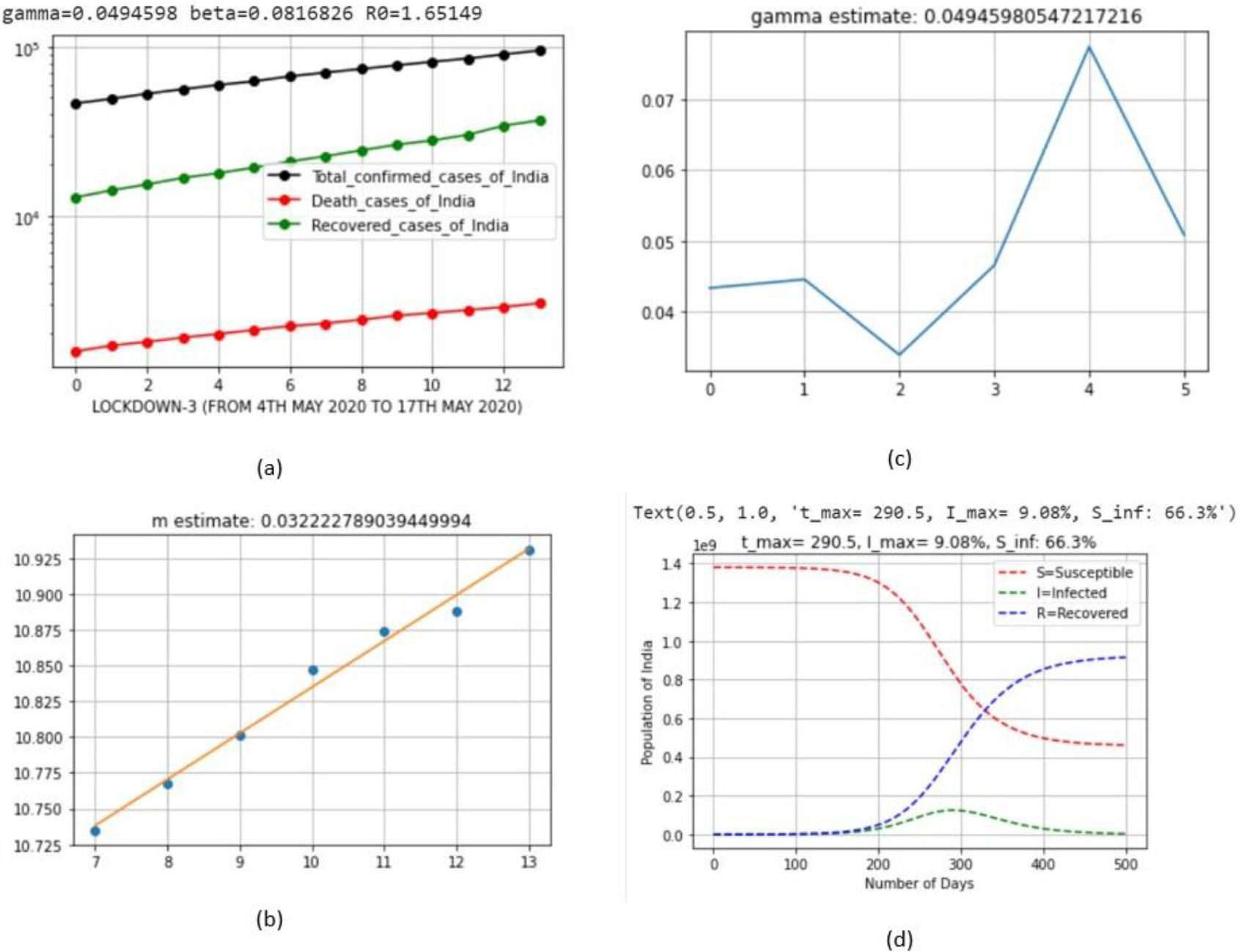
Output parameters of SIR model for lock down 3

**Figure 10:**
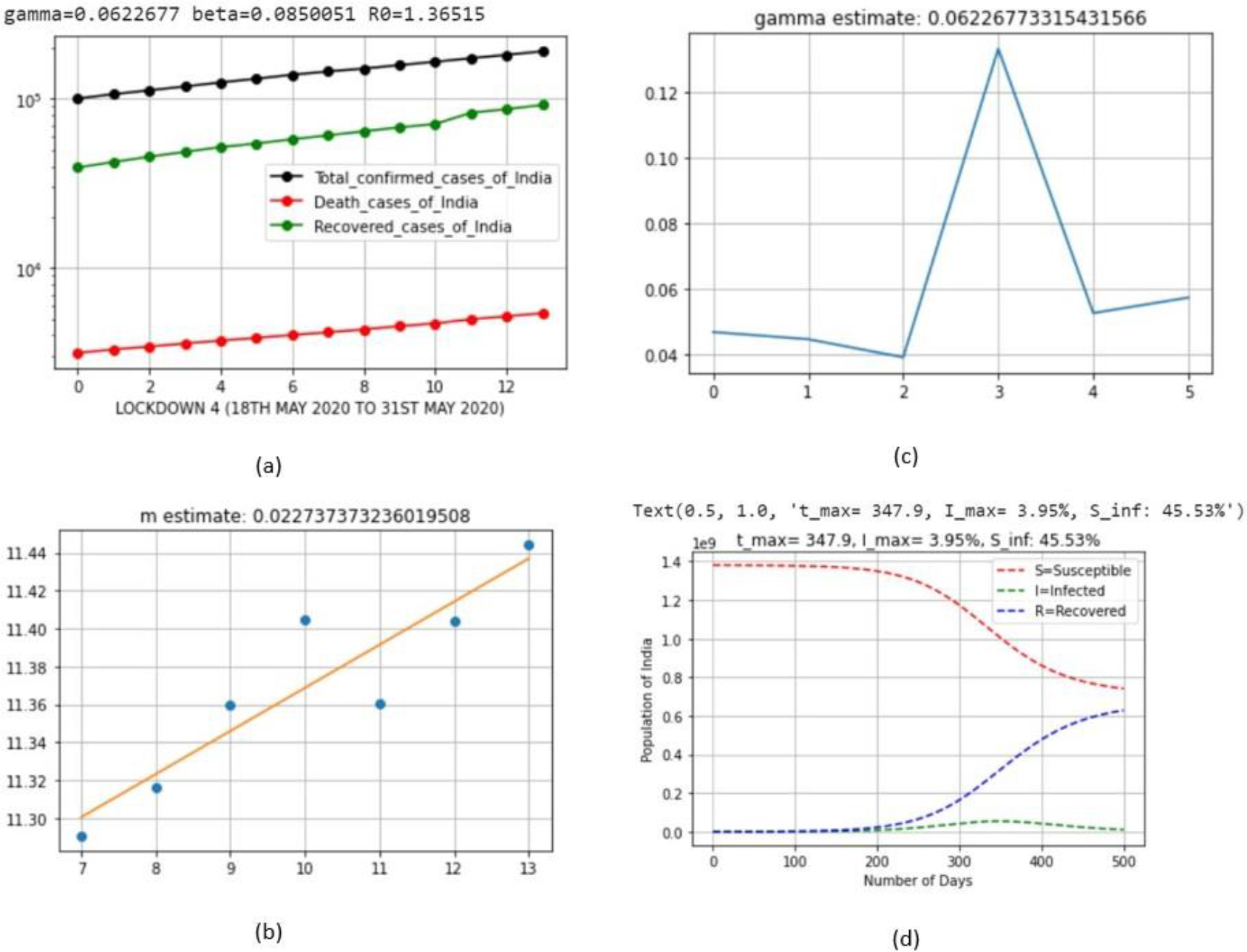
Output parameters of SIR model for lock down 4

**Figure 11:**
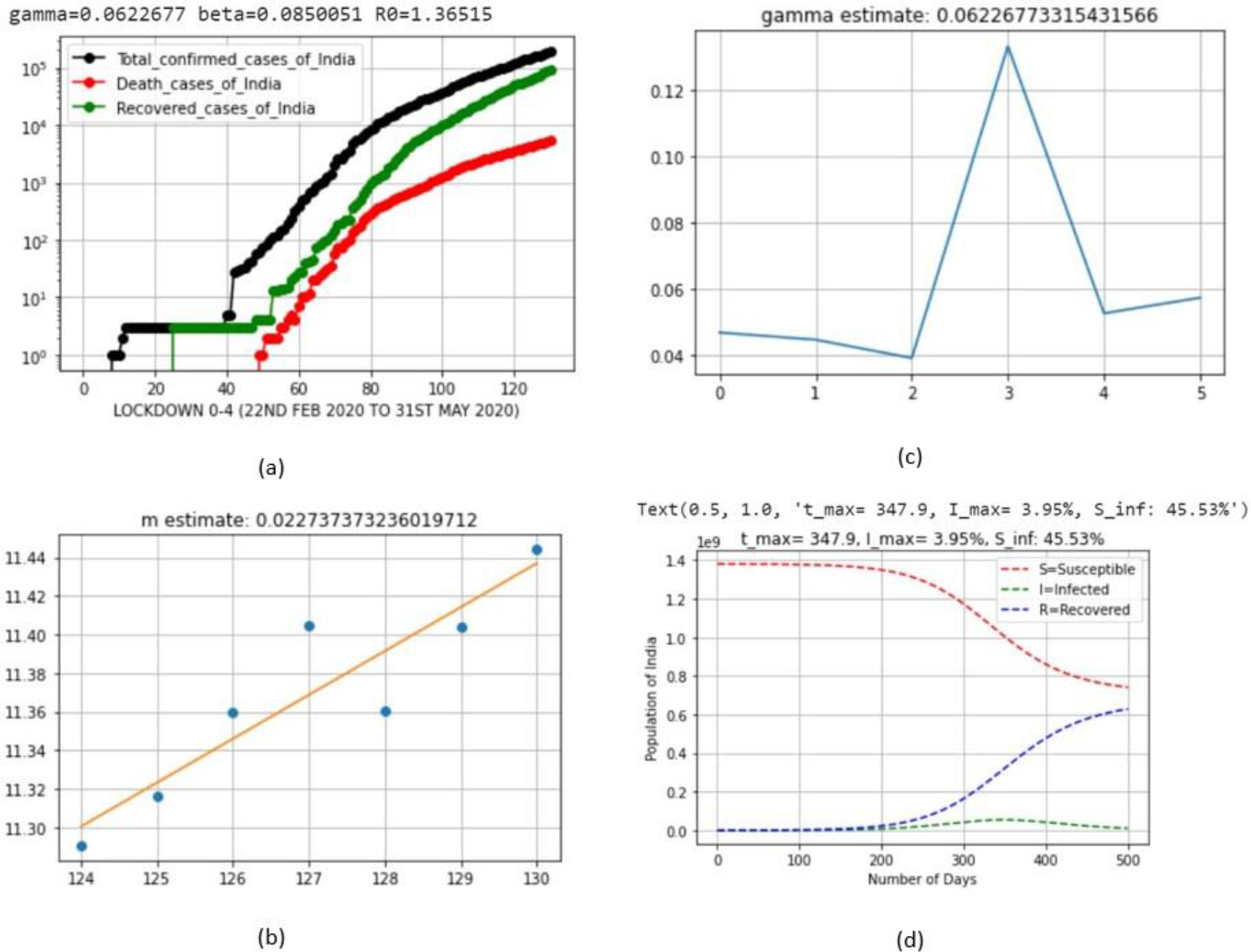
Output parameters of SIR model for lock down 0-4

## Notes

### Competing Interest Statement

The authors have declared no competing interest.

### Clinical Trial

The trial ID from an internationally recognized trial registry is not required for the mathematical case study of open online data set which are published Coronavirus COVID-19 Global Cases by the Center for Systems Science and Engineering (CSSE) at Johns Hopkins University (JHU).

### Author Declarations

The authors declare that there is no conflict of interest regarding the publication of this article.

